# Maternal age-related differences in the timing of pregnancy loss in a nationwide ART cohort

**DOI:** 10.64898/2026.09.28.26364171

**Authors:** Felix Evers, Karin Ytterberg, Alba Rocio Ramirez Jimenez, Yan Zhao, Patricia Nyeboe, Maria C Magnus, Julius Juodakis, Bo Jacobsson, Pol Solé-Navais

## Abstract

**Study question:** Are maternal age and a history of previous pregnancy losses associated with the timing of pregnancy loss for women undergoing ART treatment?

**Summary answer:** Among women who underwent ART treatment, younger women had a higher risk of very early biochemical pregnancy losses. Women with previous pregnancy losses overall experienced pregnancy losses at an earlier gestational age.

**What is known already:** Maternal age and history of pregnancy loss are two of the strongest, independent determinants of pregnancy loss risk.

**Study design, size, duration:** This retrospective cohort study comprises all embryo transfers in Sweden between 2007 and 2022 (n = 202,008). The Swedish Quality Register for Assisted Reproduction (Q-IVF) provides information on confirmed pregnancies from two weeks after embryo transfer. Additional information was retrieved from the Swedish Medical Birth Register and the National Patient Register.

**Participants/materials, setting, methods:** We studied 202,008 embryo transfers resulting in 80,673 biochemically verified pregnancies and 21,240 pregnancy losses. We assessed the associations between two exposures (maternal age and history of pregnancy loss) and four outcomes (implantation failure, pregnancy loss, the proportion of pregnancy losses occurring within the biochemical pregnancy window, and the gestational duration when the pregnancy loss was diagnosed). Fixed and mixed linear and logistic regression models were used, the latter to account for unmeasured confounding shared across pregnancies from the same mothers. Through analyses of subgroups including oocyte recipients (n = 3,558) and women undergoing treatment due to male infertility (n = 41,721), we attempted to tease out the effects of potential unmeasured confounders, such as maternal oocyte aneuploidies.

**Main results and the role of chance:** Mean maternal age was 34 years, 23 % had a history of pregnancy loss, implantation failure occurred after 60 % of embryo transfers and pregnancy loss occurred in 26 % after successful implantation.

Higher maternal age was associated with increased risk of implantation failure (OR = 1.02 per year; 95% CI = 1.02, 1.02), and pregnancy loss after successful implantation (OR = 1.05 per year; 95% CI = 1.05, 1.06). We observed no relationship between maternal age and the risk of implantation failure or pregnancy loss among women undergoing ART with donated oocytes. Among those who experienced pregnancy loss, the proportion of pregnancy losses within the biochemical window decreased with increasing maternal age (OR = 0.94 per year; 95% CI = 0.93, 0.94), a result consistent across all subgroups and regardless of oocyte origin.

The number of previous pregnancy losses was associated with a lower risk of implantation failure (OR = 0.82; 95% CI = 0.81, 0.84), but not with pregnancy loss in the index pregnancy after accounting for within-women correlations using mixed-effects models (OR = 1.02; 95% CI= 0.99, 1.04).

**Limitations, reasons for caution:** Since all women studied underwent ART treatment, the findings may not apply to naturally conceived pregnancies. The gestational duration of pregnancy losses reflects the time at diagnosis, and thus is an upper bound for the pregnancy loss.

**Wider implications of the findings:** This study confirms (oocyte) maternal age and history of pregnancy loss as influential in risk of implantation failure and pregnancy loss, and suggests that maternal age may delay the timing of the pregnancy loss, particularly among pregnancy losses occurring within the biochemical pregnancy window.

**Study funding/competing interest(s):** Funding for this research project was granted by the Swedish Research Council; the Swedish Society for Medical Research; the Agreement concerning research and education of doctors; FORTE; Swedish Research Council for Health, Working Life and Welfare; the Hjalmar Svensson Foundation; the Research Council of Norway. The authors declare no conflicts of interest.

**Trial registration number:** N/A

## Introduction

Pregnancy loss is the most common pregnancy complication; worldwide, 15 % of clinically recognized pregnancies end in early pregnancy loss, an estimated 23 million pregnancies each year^1^. The total reproductive losses are much higher, with estimates suggesting that 40-60 % of all conceptions are lost before birth^2^. The total burden of this is enormous, whether measured in terms of physical and psychological stress for the woman and others affected, or by healthcare costs. Two of the strongest risk factors for pregnancy loss are maternal age and a history of pregnancy loss^3^. The risk of pregnancy loss is slightly increased in adolescents and sharply in women above 35 years of age, and over half of all clinically recognized pregnancies end in pregnancy loss in women older than 45 years of age^4^. The risk of pregnancy loss increases by about 10 % for each additional pregnancy loss, reaching 42 % in women with three or more previous pregnancy losses^1,5^.

Aneuploidy (an abnormal number of chromosomes) or other chromosomal errors arising in meiosis are considered to be the cause of the age-related increased risk of pregnancy loss^6,7^. The meiotic errors originate in the oocyte rather than in the spermatocyte^8^, and affect all cells of the fetus. On the other hand, chromosomal errors of mitotic origin arise during embryonic/fetal development, and therefore may affect a subset of its cells. It would be reasonable to speculate that chromosomal errors of meiotic origin would lead to more severe consequences, and thus earlier embryo arrest or loss, compared to chromosomal errors of mitotic origin, due to the effect on all cells of the embryo. However, previous research shows that chromosomal errors of mitotic origin are more prevalent in embryos that are arrested in the first days after fertilization^9^. Furthermore, the prevalence of chromosomal errors of meiotic and mitotic origin is similar in pregnancy losses during the first trimester^10^.

This study aims to evaluate whether maternal age or history of pregnancy loss impacts the timing of pregnancy loss. Subanalyses in women receiving treatment with donated oocytes aid in understanding if it is the maternal or the oocyte age that is at the centre of the issue. To this end, we studied women who underwent ART treatment, using data from the Swedish National Registry of Assisted Reproduction (Q-IVF), which provides information beginning as early as two weeks following embryo transfer with follow-up throughout the whole pregnancy. We estimated the associations of maternal age and history of pregnancy loss with implantation failure, pregnancy loss among implantation successes, the proportion of pregnancy losses occurring within the biochemical pregnancy window and the gestational timing of the pregnancy loss.

## Methods

### Ethics

This study was approved by the Swedish Ethical Review Authority (Dnr 2025-09110-02), which waived patient informed consent. The National Board of Health and Welfare permitted the use of the data from the Swedish Medical Birth Register, the Q-IVF, and the National Patient Register. All individual-level data were pseudonymized.

### Study sample

We studied women registered with an IVF cycle in the Q-IVF registry^11^ between 2007 and 2022, with linked information obtained from the Swedish Medical Birth Register^12^ (1973-2022) and the Patient Register^13^ (1997 – 2022). Because maternal date of birth/age at treatment is not available in the Q-IVF, only women present in both Q-IVF and the Medical Birth Register were included. Women could be represented in the Medical Birth Register either through their own birth record (born after 1973) or as mothers of a registered delivery (live or stillborn). The Q-IVF includes nearly all fresh and frozen embryo transfer cycles performed in Sweden during the studied period. All public and private clinics report to the Q-IVF, patients are informed about it and may opt not to include their data, but this rarely occurs. From the Q-IVF, we obtained information on embryo transfer date, result of the embryo transfer (i.e., implantation failure, biochemical pregnancy loss, clinical pregnancy loss, ectopic pregnancy, elective abortion or live birth), the date of live birth or pregnancy loss, embryo culture time, fresh or frozen embryo transfer-cycle, and the use of donated oocyte or sperm. From the Patient Register, we extracted recorded diagnoses (ICD codes, **Supplementary Table 1**) for pregnancy losses prior to the index embryo transfer, female and male infertility, as well as the date when the ICD codes were registered. The ICD codes were used to count the number of previous pregnancy losses. From a total of 272,551 ART treatment cycles, we restricted the cohort to cycles in which an embryo transfer was performed, identified by the presence of an embryo transfer date, thereby excluding insemination cycles and other treatment cycles without a recorded embryo transfer (n = 63,415). Then, we excluded multiple pregnancies (n = 3,554), those where the pregnancy ended in elective abortion (n = 635), as ectopic pregnancy (n = 613), with unknown status (noted as “unknown” in the register, n = 922), unknown embryo incubation length (n = 1,389), and unknown maternal age (n = 15), rendering a total of 202,008 embryo transfer cycles included in this study.

### Exposure and outcomes

A summary of the variable definitions used is presented in **Table 1**. Maternal age at treatment was estimated based on the difference between the date of the embryo transfer from the Q-IVF and maternal date of birth from the Medical Birth Register. In oocyte recipients, maternal age referred to the age of the recipient; information on donor age was not available. The number of previous pregnancy losses was determined by registered ICD codes for pregnancy loss (any gestational duration) in the Patient Register. To account for potential overlap between two ICD codes from the same pregnancy loss, we counted a single pregnancy loss whenever two or more ICD codes tagging pregnancy loss were registered within 42 days of each other, as has been done by others^4^. The number was assessed as a count variable and as a categorical variable with the following groups: no previous pregnancy loss, one, two and three or more.

**Table 1.** Exposure and outcome definitions.

| **Term** | **Definition** |
| --- | --- |
| **Exposures** |  |
| Maternal age | Maternal age at embryo transfer |
| Number of previous pregnancy losses (count or categorical as 0/1/2/3+) | Number of previous pregnancy losses to the index pregnancy registered in the Patient Register |
| **Outcomes** |  |
| Implantation failure | Embryo transfers not resulting in a positive pregnancy test, according to Q-IVF |
| Pregnancy loss | Embryo transfers with a positive pregnancy test but not reaching live birth, according to Q-IVF |
| Biochemical pregnancy loss | Embryo transfers with "biochemical pregnancy" registered in the Q-IVF among all pregnancy losses; pregnancy loss within the biochemical pregnancy window |
| Gestational duration | Sum of number of days of embryo culture and time between date of embryo transfer and pregnancy loss as registered in the Q-IVF |

Then four outcomes were defined (**Table 1**). Implantation failure, pregnancy loss and biochemical pregnancy loss in the index pregnancy were defined using the Q-IVF. Implantation failure was defined as an embryo transfer for which there was no outcome registered (but not flagged as “unknown”). Pregnancy loss was defined as an embryo transfer with a subsequent positive pregnancy test, but no live birth. Biochemical pregnancy loss corresponds to a pregnancy loss within the biochemical pregnancy window, i.e. with a positive pregnancy test, but before positive ultrasound. The gestational duration of the pregnancy was calculated as the sum of the time (in days) of embryo culture, and time between the date of embryo transfer and the date of the end of the pregnancy (live birth or pregnancy loss).

### Statistical analysis

A summary of the analyses performed is presented in **Supplementary Table 2**. The proportion of implantation failure is the number of embryo transfers with failed implantation, divided by all embryo transfers. The proportion of pregnancy loss is the number of pregnancy losses divided by the number of embryo transfers with successful implantation.

We described the proportions of implantation failure and pregnancy loss separately for embryo transfers using autologous or donated oocytes (used as a negative control for the maternal age-linked increased risk of aneuploidies). We estimated the effects of maternal age on implantation failure and pregnancy loss using mixed effects regression models with a mother-specific random intercept to account for repeated embryo transfers. This allows us to use the full sample of transfers and to adjust for shared confounders between multiple embryo transfers in the same woman. The exposure was maternal age, and the models were additionally adjusted for the year of transfer, the number of previous pregnancy losses, the number of previous embryo transfer failures and the time of embryo incubation.

The association between maternal age and the time to pregnancy loss was assessed using two complementary measures: 1) the proportion of pregnancy losses which occurred within the biochemical pregnancy window, and 2) differences in the gestational timing of the pregnancy losses (evaluated among all pregnancy losses). For the proportion of pregnancy losses occurring within the biochemical pregnancy window, we use logistic mixed effect models where cases are defined as biochemical pregnancy losses and controls are defined as all other pregnancy losses. In these models, exposures were maternal age (continuous) or maternal age tertiles, and were adjusted for the year of transfer, the number of previous pregnancy losses and the time of embryo incubation. We explored the associations between maternal age (continuous and in tertiles) and the gestational duration when the pregnancy loss was diagnosed using mixed effects linear regression models. To test whether the effects observed were driven by the age-related increase in meiotic aneuploidies, we ran the same models in oocyte recipients, which allowed us to distinguish between the effects of maternal age vs oocyte age. Since reasons for undergoing ART treatment may differ according to maternal age, we repeated the same analysis on a group of women undergoing treatment due to male factor infertility defined as women using autologous oocytes, with a registered ICD code for male infertility, but none for female infertility. The intentions of the subgroup analyses on the oocyte recipient– and male factor infertility groups were to assess the effect of oocyte aneuploidies on the gestational duration when the pregnancy loss occurred, and to reduce confounding due to age-dependent reasons for undergoing ART treatment.

We also conducted a sensitivity analysis to address the potential selection bias introduced by linkage through the Medical Birth Register. Such a bias could occur because our data did only include women who were born in Sweden after 1973 or gave birth in Sweden after 1973. The sensitivity analysis was limited to women who were born in Sweden and had their embryo transfers after 2018. Given the small sample size, the sensitivity analysis was not performed in women using donated oocytes. In this sub-sample, we assessed the associations between maternal age and the risk of pregnancy losses which occurred within the biochemical pregnancy window and the duration of gestation in pregnancy loss (for all pregnancy losses as well as for the group having a pregnancy loss within the biochemical pregnancy window and the group having a pregnancy loss at higher gestation separately), as mentioned in points 1) and 2) above.

The associations between the number of previous pregnancy losses (count or categorized) and index pregnancy outcomes (implantation failure, pregnancy loss, biochemical pregnancy loss, gestational duration) were assessed with mixed effects models to account for multiple embryo transfers in the same woman, *de facto* adjusting for shared confounders between the different embryo transfers of each woman. These models were adjusted for maternal age (continuous), incubation days, the number of previous failed embryo transfers and year of transfer (logistic or linear regression models were used as appropriate). Because we believe that previous pregnancy loss is a factor that captures underlying materno-fetal conditions that are shared across pregnancies, rather than it being causative of subsequent losses, we repeated the analyses using fixed effect models where only one randomly selected embryo transfer per mother was used.

All analyses were conducted in R (v4.4.2, “Pile of Leaves”), with mixed effects models being implemented with *lme4*^14^ and marginal means being estimated with *emmeans*^15^. We organized our code in a Snakemake workflow^16^, which is available at https://github.com/mosaic-Wlab/maternal-age-and-pregnancy-loss.

## Results

During the period 2007-2022, and after exclusion criteria, 202,008 embryo transfers were performed in 71,715 women (**Figure 1**, **Table 2** and **Supplementary Tables 3-5**). Mean age at embryo transfer was 34 years (standard deviation, SD = 4 years), 23 % of women had a history of pregnancy loss, implantation failure occurred in 60 % of embryo transfers and pregnancy loss in 26 % after successful implantation. Embryo transfer at the cleavage stage was the most common (2-3 days, 61 %), followed by transfers at the blastocyst stage (5-6 days, 38 %). Oocyte recipients accounted for 2.2 % of embryo transfers. 51 % and 21 % of embryo transfers were due to female and male infertility, respectively.

**Figure 1.**
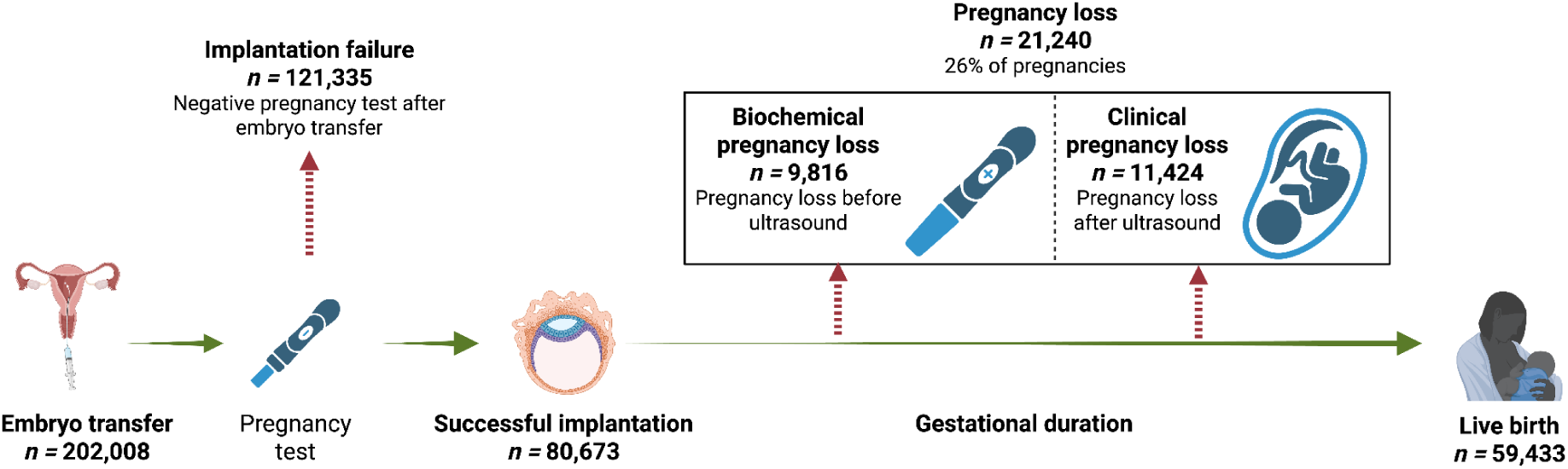
Study design and outcomes investigated in this study.

**Table 2.** Descriptive characteristics of the studied embryo transfer cycles.

|  | All<br>(n transfers = 202,008) | Implantation success<br>(n = 80,673) |
| --- | --- | --- |
| Age, years | 34.47 (4.45) | 33.91 (4.35) |
| Missing, n | 0 | 0 |
| Gestational duration, days | 214.53 (90.65) | 214.53 (90.65) |
| Missing, n | 125,468 | 0 |
| Female infertility, n | 118,139 (58.48) | 50,227 (62.26) |
| Male infertility, n | 41,721 (20.65) | 17,537 (21.74) |
| Incubation days, n |  |  |
| 1 | 68 (0.03) | 7 (0.01) |
| 2 | 95,424 (47.24) | 33,138 (41.08) |
| 3 | 27,697 (13.71) | 9,710 (12.04) |
| 4 | 1,398 (0.69) | 542 (0.67) |
| 5 | 61,094 (30.24) | 30,622 (37.96) |
| 6 | 16,295 (8.07) | 6,641 (8.23) |
| 7 | 32 (0.02) | 13 (0.02) |
| Missing, n | 0 | 0 |
| Frozen embryo transfer, n | 89,168 (44.14) | 38,310 (47.49) |
| Outcome of embryo transfer, n |  |  |
| Implantation failure | 121,335 (60.06) | – |
| Biochemical loss | 7,271(3.60) | 7,271 (9.01) |
| Fetal loss <13 gw | 12,080 (5.98) | 12,080 (14.97) |
| Fetal loss 13-22 gw | 1,434 (0.71) | 1,434 (1.78) |
| Fetal loss 22-28 gw | 130 (0.06) | 130 (0.16) |
| Fetal loss >28 gw | 325 (0.16) | 325 (0.40) |
| Liveborn | 59,433 (29.42) | 59,433 (73.67) |
| Missing | 0 | 0 |
| Oocyte recipients, n | 3,558 (1.82) | 1,580 (2.00) |
| Missing, n | 6,083 | 1,844 |
| Sperm donation, n | 10,319 (5.16) | 4,437 (5.54) |
| Missing, n | 2,079 | 576 |
| Performed treatment, n |  |  |
| ICSI | 85,671 (42.63) | 33,495 (41.71) |
| IVF | 103,771 (51.63) | 41,866 (52.14) |
| Combination, n | 11,531 (5.74) | 4,934 (6.14) |
| Missing, n | 1,035 | 378 |
| Previous pregnancy losses, n |  |  |
| 0 | 156,038 (77.24) | 62,234 (77.14) |
| 1 | 33,166 (16.42) | 13,234 (16.40) |
| 2 | 9,222 (4.57) | 3,732 (4.63) |
| 3 | 3,582(1.77) | 1,473 (1.83) |
| Missing, n | 0 | 0 |

Data are presented as mean (standard deviation) or counts (percentage). ICSI: intracytoplasmic sperm injection; IVF: In vitro-fertilization.

### Maternal age increases the risk of implantation failure and pregnancy loss

We estimated the risk of implantation failure and pregnancy loss by maternal age in a population of women undergoing ART treatment using autologous oocytes (**Figure 2A**). Perhaps unsurprisingly, maternal age was associated with an increased risk of implantation failure (logistic mixed effects models, OR 1.02 per year; 95% CI 1.02, 1.02; p-value = 6.63 x 10^-64^; n cases = 115,118; n controls = 77,249) and pregnancy loss (logistic mixed effects models, OR 1.05 per year; 95% CI 1.05, 1.06; p-value = 1.07 x 10^-139^; n cases = 20,347; n controls = 56,902). The highest risk of pregnancy loss was observed in women aged 44 years or older (0.79; 95% CI 0.76, 0.81; n = 171) and the lowest in women aged 20 years or younger (0.13; 95% CI 0.07, 0.19; n = 40), without noticeable differences until 30 years of age (**Figure 2A** and **Supplementary Table 6**). This contrasts with the proportions of pregnancy loss previously reported in women after natural conception, where the lowest proportion was observed in women aged 24-29 years old^4^. These associations remained consistent when restricting the sample to Swedish-born women undergoing embryo transfer after 2018, reducing the potential concern that the findings were driven by sample selection bias related to linkage through the Medical Birth Register (**Supplementary Table 7**).

**Figure 2.**
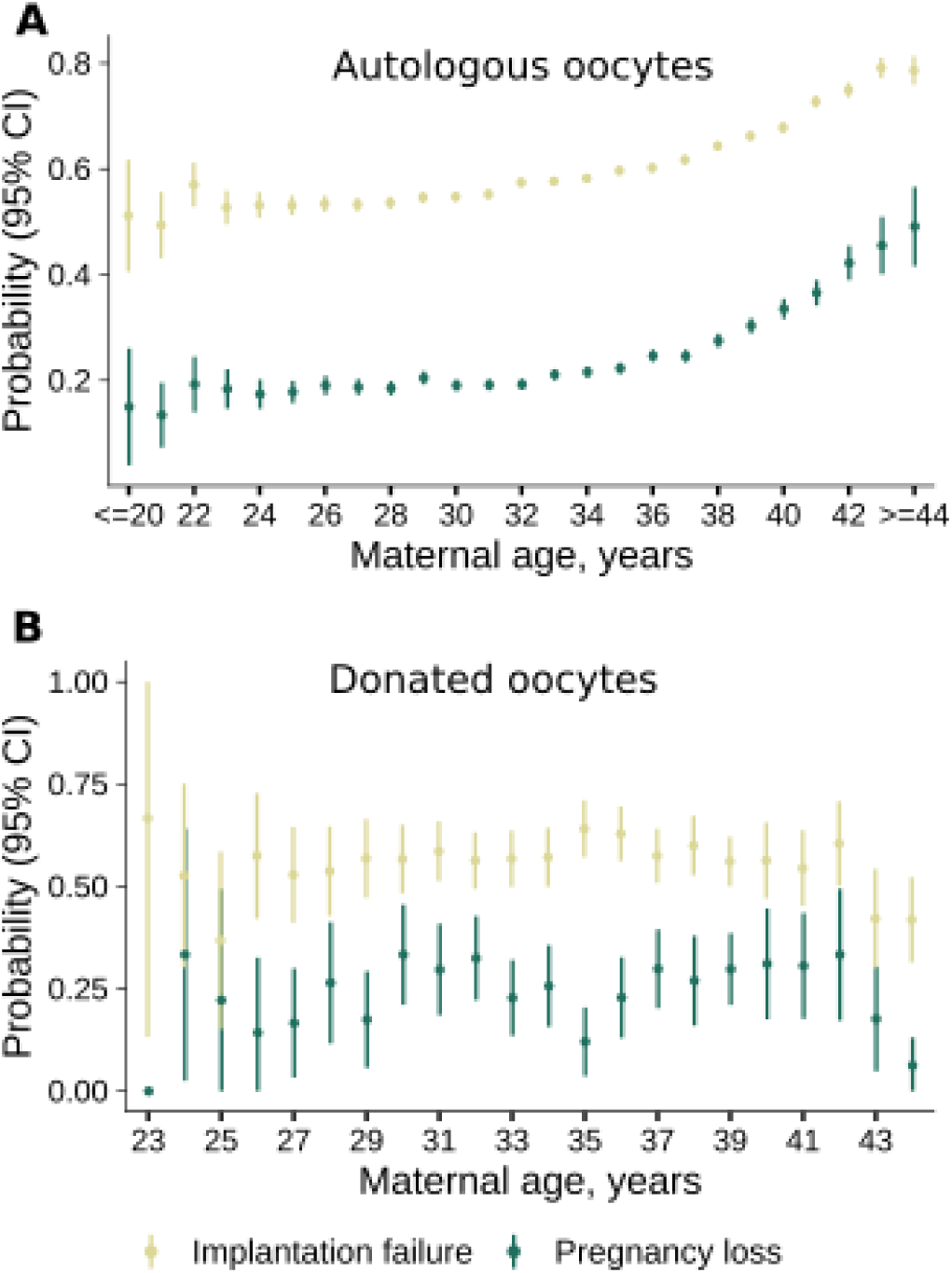
Proportions of implantation failure (yellow) and pregnancy loss (green) according to maternal age in women undergoing ART treatment. Panel A shows the proportions in cycles where autologous oocytes were used (n = 192,977 and n = 73,256 for implantation failure and pregnancy loss, respectively), and Panel B shows the proportions in oocyte recipients (n = 2,711 and n = 1,169 for implantation failure and pregnancy loss, respectively). Vertical bars represent 95% CI. The proportion of pregnancy loss was estimated in embryo transfer cycles with a successful implantation.

We considered oocyte recipients a negative control group, to test whether oocyte age is associated with the risks of implantation failure or pregnancy loss (**Figure 2B** and **Supplementary Table 8**). We observed no association between the recipient’s age and the risk of implantation failure (OR = 1.02; 95% CI = 0.99, 1.04; p-value = 0.094; n cases = 1,542; n controls = 1,169, n individuals = 1,237) or the risk of pregnancy loss (OR = 0.99; 95% CI = 0.99, 1.02; p-value = 0.576; n cases = 358; n controls = 811, n individuals = 904). This suggests that the age-related risk increase of both implantation failure and pregnancy loss is likely a result of higher aneuploidy proportions in older oocytes, as has been previously shown ^8,10,17^.

### The proportion of pregnancy losses within the biochemical pregnancy window is higher in younger women

To assess whether maternal age was associated with earlier pregnancy losses, we used logistic mixed effects models to regress maternal age on the proportion of pregnancy loss within the biochemical pregnancy window. Given the marked role of maternal age in embryos conceived with autologous oocytes compared to donated oocytes, we conducted subsequent analyses separating these into two groups: treatments with autologous oocytes (n pregnancy loss = 20,347) and treatments with donated oocytes (n pregnancy loss = 893). Increasing maternal age was associated with a lower proportion of pregnancy loss within the biochemical pregnancy window (**Figure 3**, OR = 0.94 per year increase; 95% CI = 0.93, 0.94; p-value = 1.0 x 10^-70^; n cases = 3,732; n controls = 7,064). Women in the third tertile of maternal age had a 47% lower risk of pregnancy loss within the biochemical pregnancy window compared to women in the first tertile (**Supplementary Figure 1**, OR = 0.53; 95% CI = 0.49, 0.57; p-value = 3.1 x 10^-59^), suggesting that pregnancy losses tended to occur later in older women. The association was also observed in women undergoing treatment due to male factor infertility (**Supplementary Figure 2**, OR = 0.95 per year increase; 95% CI = 0.92, 0.98; p-value = 7.1 x 10^-4^; n cases = 427; n controls = 885). To understand whether the observed association could be related to an increased proportion of aneuploidies in older women, the same analysis was performed in oocyte recipients. While the sample size was much lower, the effect size magnitude remained consistent with the results obtained in women undergoing ART using autologous oocytes, although the association was not significant (**Figure 3**, OR = 0.95 per year increase; 95% CI = 0.90, 1.01; p-value = 0.09; n cases = 128; n controls = 230; n individuals = 290).

**Figure 3.**
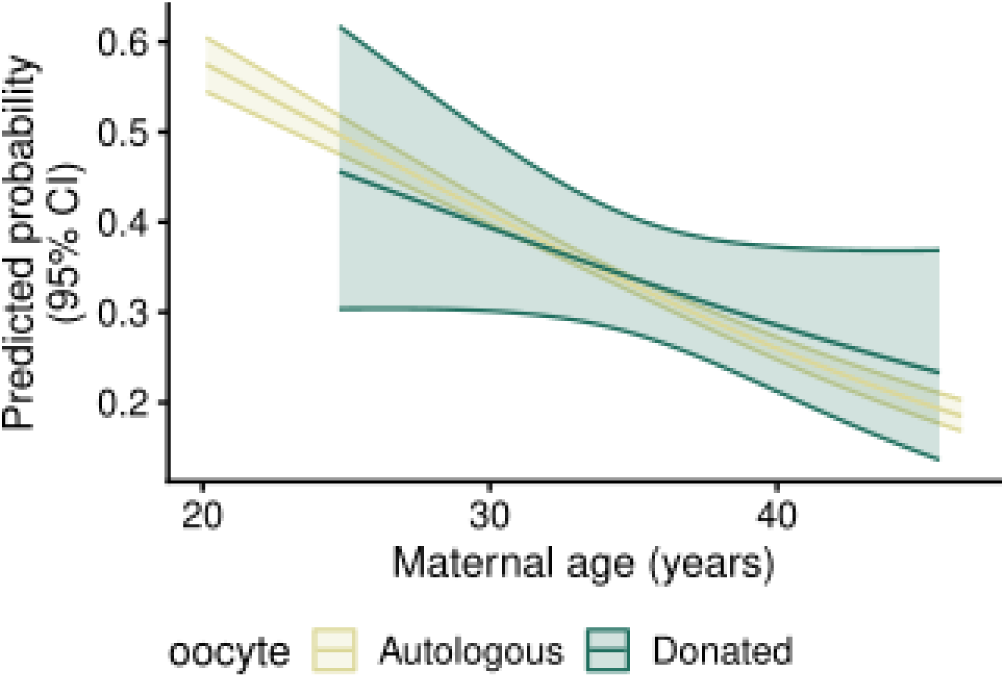
Predicted proportion of pregnancy losses that occurs in the biochemical window according to maternal age, with its 95% CI shaded. Marginal means were estimated from a logistic mixed effects model in women undergoing treatment using autologous oocytes (n cases = 3,732; n controls = 7,064) or donated oocytes (n cases = 128; n controls = 230), separately.

The effect magnitude and direction was consistent when we restricted to Swedish born women who had their embryo transfer after 2018, with a reduction in the proportion of pregnancy losses within the biochemical window (**Supplementary Table 7**, OR = 0.93; 95% CI = 0.92, 0.94; p-value = 4.9 x 10^-25^; n cases = 1,925; n controls = 3,153).

Given the lower proportion of pregnancy losses that occurred within the biochemical pregnancy window, we examined whether this translated into a longer gestational duration using linear mixed models (**Figure 4A, Supplementary Table 9**). In women undergoing treatment with autologous oocytes resulting in a pregnancy loss, maternal age was associated with longer duration of gestation (beta = 0.14 days; 95% CI = 0.09, 0.30; p-value = 3.0 x 10^-3^; n = 16,844; n individuals = 13,630). This positive effect appeared to primarily reflect the higher rate of biochemical pregnancy losses (beta = 0.16 days; 95% CI = 0.02, 0.30; p-value = 2.8 x 10^-2^; n = 4,113; n individuals = 3,687), while the association between maternal age and gestational duration was negative in clinical pregnancy losses (beta = –0.22 days; 95% CI = –0.32, –0.11; p-value = 9.0 x 10^-5^; n = 12,731; n individuals = 10,774), similar to what is observed in live-borns^18,19^. We also observed longer gestations at pregnancy loss in women undergoing ART due to male-factor infertility (beta = 0.43 days; 95% CI = 0.01, 0.85; p-value = 0.046; n = 917; n individuals = 758; **Supplementary Figure 3**) and using donated oocytes (beta = 0.56 days; 95% CI = –0.08, 0.1.20; p-value = 0.090; n = 275; n individuals = 238; **Figure 4B**). However, the sample sizes of these subgroups are small and confidence intervals wide, warranting caution in their interpretation. The small sample size also hindered us from further sub-sampling into biochemical and clinical pregnancy losses.

**Figure 4.**
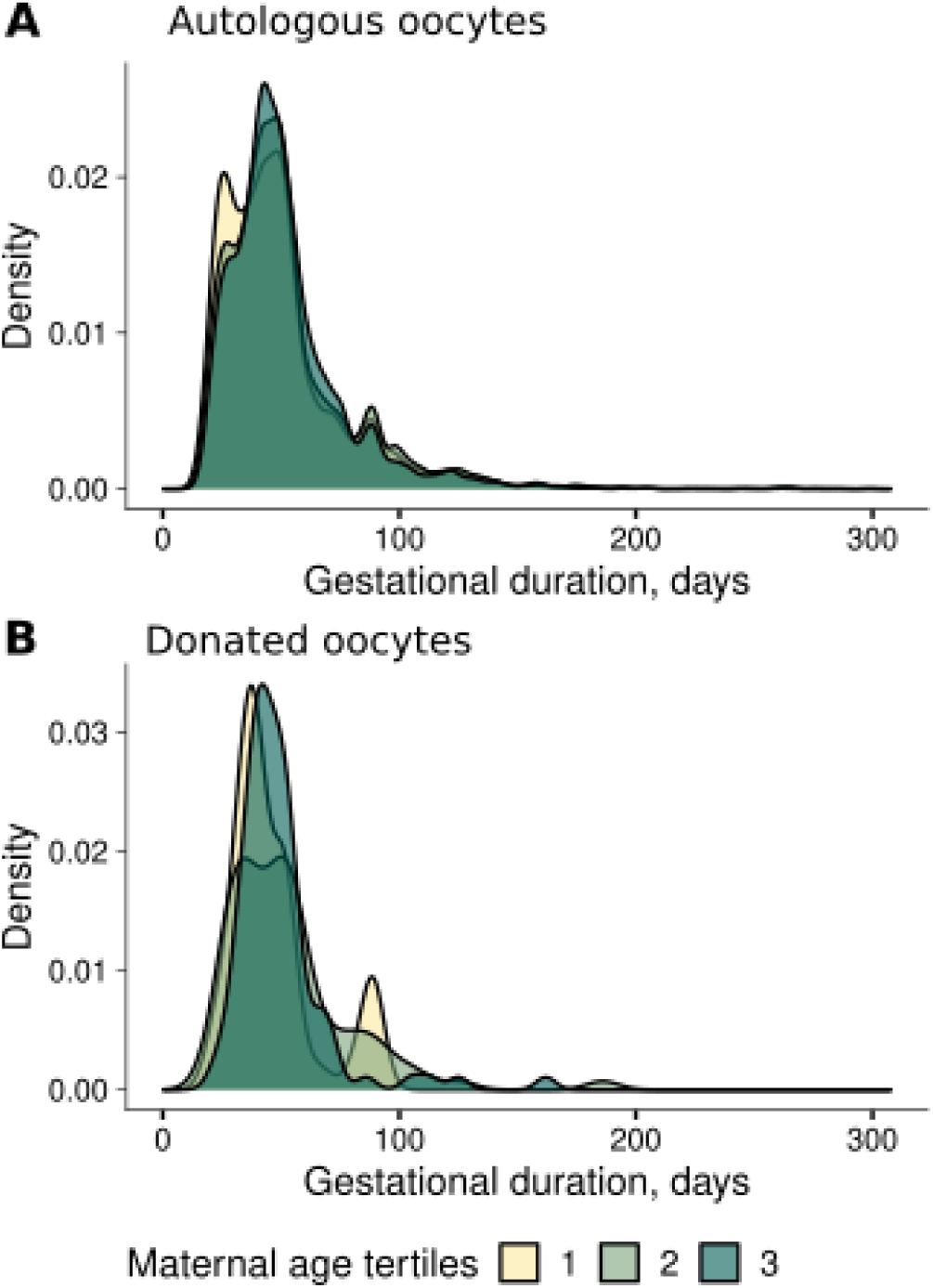
Distribution of gestational duration (days) in pregnancy losses according to maternal age tertiles in women using autologous (A) or donated (B) oocytes. Sample sizes for maternal age tertiles in women with autologous oocytes (A) were 5,803, 6,301 and 8,243 for the first (<31.9 years old), second (>=31.9 and <36.0 years old) and third (<48.0 years old) tertile, respectively. Sample sizes for maternal age tertiles in women with donated oocytes (B) were 131, 110 and 117 for the first (<33.6 years old), second (>=33.6 and <37.6 years old) and third (<45.6 years old) tertile, respectively.

In the corresponding sensitivity analyses of gestational duration, the association remained positive when all pregnancy losses were considered, but the effect estimates were larger (**Supplementary Table 7**, beta = 0.48 days; 95% CI = 0.30, 0.65; p-value = 6.3 x 10^-8^; n = 4,410; n individuals = 3,733). Among biochemical pregnancy losses, the estimates were similar in direction and effect compared to the main analyses (beta = 0.22 days; 95% CI = –0.03, 0.46; p-value = 0.085; n = 1,387; n individuals = 1,261). In clinical pregnancy losses the estimate was in the opposite direction compared to the main analyses (beta = 0.15 days; 95% CI = –0.05, 0.36; p-value = 0.162; n = 3,023; n individuals = 2,685), but with wide confidence intervals.

**History of pregnancy loss is not associated with the proportion of pregnancy losses occurring within the biochemical pregnancy window or the gestational duration when pregnancy loss is diagnosed**

We observed that the greater the number of previous pregnancy losses, the lower the risk of implantation failure (**Supplementary Figure 4**, OR = 0.82; 95% CI = 0.81, 0.84; p-value = 6.14 x 10^-133^; n cases = 115,118; n controls = 77,249; n individuals = 68,661). We recognize that modelling the association between previous pregnancy losses and implantation failure is not trivial due to the correlation between the number of previous losses (which obviously happened after a successful implantation) and implantation success in the index pregnancy. Using logistic mixed effects models, we didn’t observe an association between the number of previous pregnancy losses and pregnancy loss in the index pregnancy (**Supplementary Figure 4**, OR = 1.02; 95% CI= 0.99, 1.04; p-value = 0.129; n cases = 20,347; n controls = 75,215; n individuals = 53,153). By using mixed effects models, the effects of unmeasured confounding factors that are shared across pregnancies are adjusted for. Using fixed effects linear models (selecting a random pregnancy from each mother), we observe a positive association between previous pregnancy losses and subsequent ones (OR = 1.10; 95% CI = 1.07, 1.14; p-value = 1.14 x 10^-8^; n cases = 11,177; n controls = 41,976; n individuals = 53,153).

Among women who experienced a pregnancy loss, we assessed the association between the number of previous pregnancy losses and subsequent proportion of pregnancy losses occurring within the biochemical pregnancy window restricted to those undergoing treatment using autologous oocytes. Using logistic mixed effects regression models, we observed no association (**Table 3**, OR = 1.02; 95% CI = 0.97, 1.06; p-value = 0.465; n cases = 6,983; n controls = 13,364; n observations = 20,347; n individuals = 15,953). We observed an association between the number of previous pregnancy losses and gestational timing of the losses, indicating a tendency for an earlier timing of the loss with increasing number of previous losses with borderline significance (**Table 3**, beta = –0.58; 95% CI = –1.12, –0.04; p-value = 0.035; n individuals = 13,630; n observations = 16,844).

**Table 3.** Associations between the number of previous pregnancy losses and the proportion of pregnancy losses occurring within the biochemical pregnancy window and gestational duration at pregnancy loss diagnosis.

|  | Biochemical pregnancy loss |  | Gestational duration at pregnancy loss diagnosis |  |
| --- | --- | --- | --- | --- |
|  | OR (95% CI) | p-value | Beta (95% CI) | p-value |
|  | n individuals = 15,953 |  | n individuals = 13,630 |  |
| Mixed model | 1.02 (0.97, 1.06)<sup>a</sup> | 0.456 | -0.58 (-1.12, -0.04)<sup>b</sup> | 0.035 |
| Standard model | 0.99 (0.93, 1.05)<sup>c</sup> | 0.798 | -0.65 (-1.49, 0.18)<sup>d</sup> | 0.126 |

Estimates were obtained using logistic or linear mixed effects models for biochemical pregnancy loss and gestational duration, respectively. Only embryo transfer cycles with autologous oocytes leading to pregnancy loss were included. The models were adjusted for maternal age, year of transfer and embryo incubation length.
<sup>a</sup> n cases = 6,983, n controls = 13,364
<sup>b</sup> n = 16,844
<sup>c</sup> n cases = 3,717, n controls = 7,398
<sup>d</sup> n = 9,176

## Discussion

In this register-based study, we evaluated maternal age and history of pregnancy loss and their association with the timing of pregnancy loss in a population undergoing ART. We observe a higher risk of pregnancy loss with increasing maternal age, as previously shown^4^. By using oocyte recipients as negative controls, our results support that oocyte age, rather than maternal age, likely drives the increased risk, since no such risk is seen in oocyte recipients. Higher maternal age was associated with increased risk of implantation failure and pregnancy loss by 5-6 % per year, which aligns with previous studies^4,20,21^. Higher maternal age was associated with a lower proportion of pregnancy losses occurring within the biochemical pregnancy window and, among biochemical pregnancy losses, with a longer gestational duration at diagnosis.

The negative association between the number of previous pregnancy losses and subsequent implantation failure is intriguing, biologically as well as statistically. We suggest that the effect of previous pregnancy losses on subsequent ones is driven by factors that are shared across pregnancies, and is not causal *per se*. The hypothesis that women with more pregnancy losses tend to have higher endometrial receptivity and lower selectivity ^6,21,22^ fits our results, with women having more pregnancy losses having lower odds of implantation failure and higher risk of pregnancy loss in subsequent embryo transfers. However, it is also true that women with previous pregnancy losses by definition have had previous implantation success. Studying women with recurrent pregnancy loss in an ART treatment context will create a scenario where the magnitude of the observed effect likely will be more extreme than it would be in a corresponding cohort conceiving naturally.

The analysis of the association between previous pregnancy loss and the risk of subsequent pregnancy loss is particularly interesting from a methodological point of view, where the use of the logistic mixed effects model shows no such association while standard logistic regression does. This is coherent with the experience from clinical practice, where a great proportion of investigations of women with recurrent pregnancy loss show no obvious cause, while the clinical pattern on the individual level remains clear. The logistic mixed-effects model adjusts for the individual factor that is shared across transfers, and that is unadjusted for in the fixed-effects logistic regression model. Our interpretation of the result is that it is the underlying individual’s causes, rather than the previous pregnancy losses themselves, that account for the increased risk for pregnancy loss for these individuals.

Surprisingly, older women had nearly half the risk of having a pregnancy loss within the biochemical pregnancy window compared with younger women. This pattern was also observed among women undergoing ART for male-factor infertility and among those using donated oocytes. The association was essentially unchanged in a sensitivity analysis restricted to Swedish-born women undergoing embryo transfer from 2018 onwards, reducing the potential concerns that this finding resulted from selection bias introduced through the linkage of the Q-IVF with the Medical Birth Register. Analyses on gestational duration were not robust to sample selection bias, and thus the results regarding gestational duration should be interpreted with caution. We speculate that the negative association we observe between maternal age and the proportion of pregnancy losses occurring within the biochemical pregnancy window might be driven by the symptoms and the recognition of the pregnancy loss. This speculation is somewhat supported by the fact that the increase in gestational duration was only observed in biochemical pregnancy loss, and a negative effect of maternal age on the duration of gestation was observed in clinical pregnancy losses. However, these associations with gestational duration were not robust in sensitivity analyses and should therefore be interpreted cautiously. Pregnancy loss has a broad array of clinical presentations, from completely asymptomatic to dramatic bleeding. Since the underlying mechanisms of pregnancy loss in general are poorly understood, we must consider the possibility that differences other than strict biological ones between the age groups may steer the clinical presentation of the pregnancy loss in one direction or the other, and thus affect the time when the diagnosis is made. One such example would be if patients of a certain age group have a tendency to seek care earlier, or if the medical advice provided to the patient differs between age groups.

The strengths of this study are the number of included embryo transfers, the reliability of the data, the availability of multiple transfers for most women, the nationwide coverage over a large period in time, and its great detail in early gestational weeks. Limitations include the variations in the clinical presentations of pregnancy losses and its consequence for the documented timing of pregnancy loss, the relatively small size of the oocyte-recipient group and the lack of information about the age of the oocyte donor. Another limitation is a potential selection bias of women included in the study. By conducting sensitivity analyses, we observed that most of our results were robust to such selection bias.

To this day, preimplantation genetic testing for aneuploidies as a screening tool is not allowed in the ART clinics in Sweden. Since there is therefore no knowledge of the genetic status of the embryos in the early gestational period, embryos with aneuploidy will be transferred to a much greater extent in this medicolegal context than where the practice is allowed. The increasingly popular clinical practice of only transferring embryos at the blastocyst stage would reduce the amount of embryo transfers with aneuploid embryos. In this study, transfer at the cleavage stage was the most common, and this should in the context of this studied research subject be considered a strength, as it better allows for conclusions about aneuploidy and its impact on the timing of pregnancy loss. While we acknowledge the larger inclusion of transferred aneuploid embryos as a strength, embryos eligible for transfer are still selected based on morphology, which tracks closely with aneuploidy proportions. The discrepancy between today’s growing practice of blastocyst transfer and our studied cohort should therefore not be considered a methodological limitation per se, but it should be kept in mind when interpreting the findings of this study.

To conclude, in a Swedish ART population, we confirm that maternal age increases the risk of implantation failure, and attribute this effect specifically to oocyte ageing. The timing of the pregnancy loss appears to be later in older mothers, particularly among biochemical pregnancy losses. This counterintuitive finding warrants exploration in different cohorts and study designs.

## Data availability

Due to restrictions by national law and EU regulations, personal registry data cannot be shared publicly. Applicants seeking to access the data must apply to the Swedish Ethics Committee and the register authorities.

## Acknowledgements

We thank all ART clinics in Sweden for rigorously collecting data relevant to this study.

## Authors’ roles

All authors made substantial contributions to the project’s conception and design, as well as interpretation of the data, revised the manuscript critically, approved the final version for publication and agreed to be accountable for all aspects of the work. F.E., J.J and P.S.N. conducted data curation, visualization and analysis, as well as developing methodology and writing the original draft. A.R.R.J. and Y.Z. conducted data curation. F.E., B.J. and P.S.N. acquired funding for the project, and B.J., J.J, M.C.M. and P.S.N. supervised the project. K.Y., P.N., F.E., B.J. and P.S.N. obtained the linked register data for the project.

