## Supplementary Figures for "Maternal age-related differences in the timing of pregnancy loss in a nationwide ART cohort"

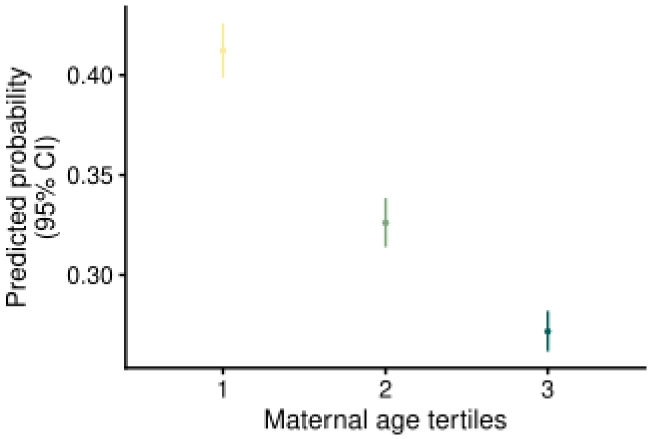


**Supplementary Figure 1.** Predicted probability of biochemical pregnancy loss (among all losses) according to maternal age tertiles in women undergoing ART with autologous oocytes.

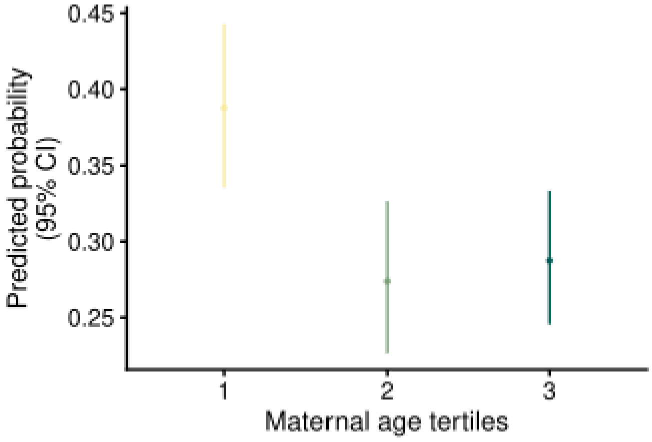


**Supplementary Figure 2.** Predicted probability of biochemical pregnancy loss (among all losses) according to maternal age tertiles in women undergoing ART because of male-factor infertility using autologous oocytes.

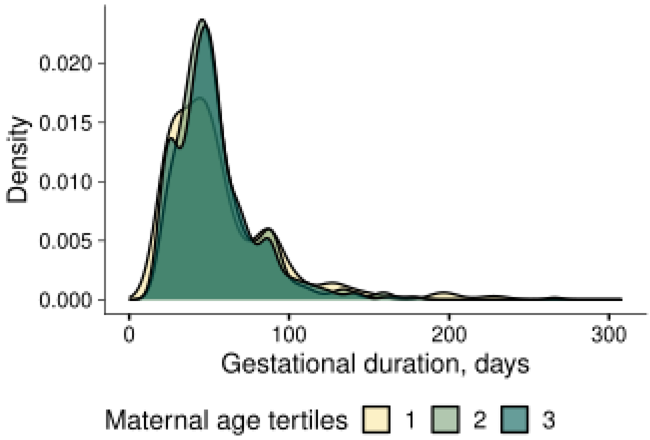


**Supplementary Figure 3.** Distribution of gestational duration in pregnancy losses according to maternal age tertiles in women using autologous with male factor infertility. Sample sizes for maternal age tertiles were 375, 393 and 544 for the first (<31.1 years old), second (>=31.1 and <35.1 years old) and third (<45.7 years old) tertile, respectively.

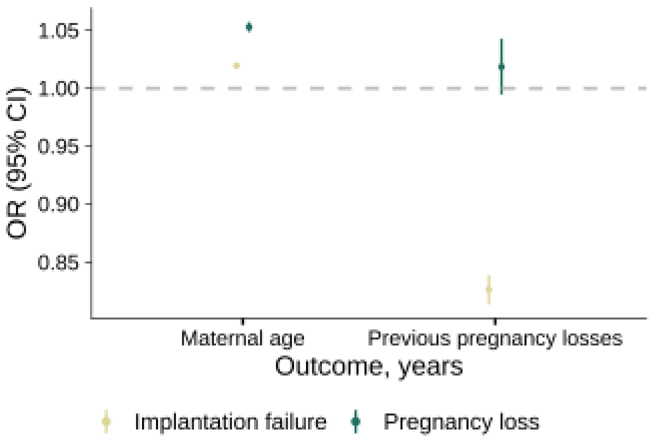


**Supplementary Figure 4.** Associations between maternal age and previous pregnancy losses on implantation failure and pregnancy loss in a sample of women undergoing ART with autologous oocytes. OR and 95% confidence intervals were estimated using mixed effects logistic regression models.
